# Latent Factors of Language Disturbance and Relationships to Quantitative Speech Features

**DOI:** 10.1101/2022.03.31.22273263

**Authors:** Sunny X. Tang, Katrin Hänsel, Yan Cong, Amir H. Nikzad, Aarush Mehta, Sunghye Cho, Sarah Berretta, Leily Behbehani, Sameer Pradhan, Majnu John, Mark Y. Liberman

## Abstract

**Background and Hypothesis:** Quantitative acoustic and textual measures derived from speech (“speech features”) may provide valuable biomarkers for psychiatric disorders, particularly schizophrenia spectrum disorders (SSD). We sought to identify cross-diagnostic latent factors for speech disturbance with relevance for SSD and computational modeling.

**Study Design:** Clinical ratings for speech disturbance were generated across 14 items for a cross-diagnostic sample (N=343), including SSD (n=97). Speech features were quantified using an automated pipeline for brief recorded samples of free-speech. Factor models for the clinical ratings were generated using exploratory factor analysis, then tested with confirmatory factor analysis in the cross-diagnostic and SSD groups. Relationships among factor scores, speech features and other clinical characteristics were examined using network analysis.

**Study Results:** We found a 3-factor model with good fit in the cross-diagnostic group and acceptable fit for the SSD subsample. The model identifies an *impaired expressivity* factor and two interrelated disorganized factors for *inefficient* and *incoherent* speech. Incoherent speech was specific to psychosis groups, while inefficient speech and impaired expressivity showed intermediate effects in people with nonpsychotic disorders. Network analysis showed that the factors had distinct relationships with speech features, and that the patterns were different in the cross-diagnostic versus SSD groups.

**Conclusions:** We report a cross-diagnostic 3-factor model for speech disturbance which is supported by good statistical measures, intuitive, applicable to SSD, and relatable to linguistic theories. It provides a valuable framework for understanding speech disturbance and appropriate targets for modeling with quantitative speech features.

## Introduction

Quantitative features derived from speech are increasingly recognized as valuable predictors and objective biomarkers for psychiatric disorders, notably including schizophrenia spectrum disorders (SSD).^1–4^ In this paper, we identify cross-diagnostic latent factors for language disturbance, then demonstrate the relevance of these factors for computational linguistic modeling in relation to general psychopathology and in SSD, particularly. Here, we regard speech as speech, without making inferences regarding “thought disorder,” a related construct which infers disruptions to thought based on observable changes in speech.

A range of speech features appear promising as predictors of psychiatric diagnoses and biomarkers of individual symptom dimensions. We use the term “speech features” broadly to indicate quantitative metrics derived from speech samples, including phonetic, acoustic and textual measures. Much of the work on speech biomarkers has been done in SSD. For example, SSD diagnosis can be classified with >80% accuracy relative to healthy volunteers (HV) using measurements of semantic distance, which quantify the “closeness” of the meaning in successive sentences or segments of words.^5,6^ This strategy can be combined with other automated measurements of syntax/parts-of-speech,^7–9^ referential ambiguity,^9^ and metaphoricity.^10^ Acoustic features (pitch, voice quality, pauses) also successfully predict SSD diagnosis.^11^ Additionally, transition to psychosis among individuals at clinical high risk can be predicted with 83-97% accuracy using automatic measurements of semantic density,^12^ detection of metaphorical speech or non-standard meanings,^10^ and a combination of semantic distance and parts-of-speech.^13,14^ Speech features are also promising biomarkers for other clinical contexts, including mania,^15^ depression,^16^ autism,^17^ and dementia.^18,19^

Presumably, speech features predict clinical characteristics by reflecting speech-related symptoms such as decreased prosody, tangentiality, and changes in quantity. However, most studies have used speech features to directly predict diagnosis or outcomes, without relating features to observable speech disturbances.^5–14^ There are weaknesses to this approach. First, there is substantial heterogeneity in speech phenotypes within diagnoses. Cross-diagnostically, and within SSD, positive/disorganized versus negative/impoverished dimensions have been consistently reported;^2,20,21^ they are poorly and sometimes even negatively correlated with one another.^22^ Therefore, greater precision may be achieved by modeling specific types of speech disturbance, rather than diagnostic phenotypes as a whole.^22–24^ Second, speech disturbances may be shared across disorders, with implications for underlying neurobiology. We need to define cross-diagnostically valid constructs in order to determine whether speech features can be used consistently across disorders as biomarkers for particular speech phenotypes. Notably, in a large sample (N=1,071), Stein et al. found three factors of speech disturbance (emptiness, disorganization, and incoherence) which were linked to changes in distinct brain regions and valid across diagnoses.^25^ Speech graph features have been shown to be a good marker of thought disorder in mania and SSD,^15^ and to be related to cross-diagnostic psychopathological dimensions.^26^ However, there are few studies examining whether quantitative speech features are consistently related to cross-diagnostic dimensions of speech disturbance.

Previous studies have reported factor analyses on rating scales for thought disorder, but results are varied, mostly limited to SSD cohorts, and relevance for computational modeling is unclear. The two-factor model distinguishing impoverished speech (e.g., poverty of speech, latency, concreteness) from disorganized speech (e.g., tangentiality, derailment, incoherence, neologisms, clanging) is the most consistent.^2,20,21^ However, “disorganized speech” is inconsistently defined across studies and encompasses a broad collection of individual symptoms which may require different computational strategies. A 6-factor model has been replicated in SSD but several factors are not likely to be relevant cross-diagnostically (e.g., idiosyncratic: word approximations and stilted speech; referential: echolalia and self-reference).^21,27^

The objective of this study was to delineate dimensions of observable speech disturbance suitable as targets for computational modeling in SSD and across diagnoses. First, we used factor analyses to identify and test interpretable models. Then, we evaluated the clinical relevance of the factor scores by comparing severity across diagnostic groups, with the expectation that clinically relevant factors should show different patterns in different groups. Finally, using network analysis, we related the factors to multi-modal speech features cross-diagnostically and within an SSD subgroup.

## Methods

### Participants

The cross-diagnostic sample included brief recordings of free speech (∼1.5-3 minutes) from 343 individuals, including 97 definitively diagnosed with SSD, 31 with affective psychosis or probable but unconfirmed psychotic disorder (PSY), 130 with other non-psychotic disorders (OD), and 76 HV (Table 1). Other psychiatric disorders included unipolar and bipolar mood disorders, anxiety disorders, OCD, borderline personality disorder, ADHD, and substance use disorders. Many participants were diagnosed with multiple comorbid conditions, so diagnoses were not mutually exclusive. Individuals with developmental, neurological, or medical conditions likely to impact speech were excluded, including intellectual disability, autism spectrum disorder, and dementia. All recruited participants signed informed consent, and human subjects research ethical approval was given by the IRB at the Feinstein Institutes for Medical Research. Additional details on ascertainment and sample collection are provided in the supplement.

**Table 1.** Participant and Sample Characteristics.

|  | Healthy<br>Volunteers (HV) | Other<br>Psychiatric<br>Disorders (OD) | Other or<br>Undetermined<br>Psychosis (PSY) | Schizophrenia<br>Spectrum<br>Disorders (SSD) | Total<br>Sample | P - Value |
| --- | --- | --- | --- | --- | --- | --- |
| **N** | 76 | 130 | 31 | 97 | 334 |  |
| **Age - mean (SD)** | 29.0 (6.7) | 23.8 (5.5) | 26.7 (6.9) | 26.8 (6.7) | 26.2 (6.6) | <0.001 |
| **Gender - n (%)** |  |  |  |  |  | 0.004 |
| **Man** | 35 (46%) | 44 (34%) | 19 (61%) | 57 (59%) | 155 (46%) |  |
| **Non-Binary** | 2 (3%) | 12 (9%) | 0 (0%) | 4 (4%) | 18 (5%) |  |
| **Woman** | 39 (51%) | 73 (56%) | 12 (39%) | 34 (35%) | 158 (47%) |  |
| **Race – n (%)** |  |  |  |  |  | 0.002 |
| **Asian** | 11 (15%) | 14 (13%) | - | 15 (16%) | 40 (14%) |  |
| **Black** | 29 (38%) | 21 (19%) | - | 41 (42%) | 91 (32%) |  |
| **Multiple** | 5 (7%) | 8 (7%) | - | 6 (6%) | 19 (7%) |  |
| **Other** | 2 (2%) | 8 (7%) | - | 14 (14%) | 25 (9%) |  |
| **White** | 29 (38%) | 60 (54%) | - | 21 (22%) | 112 (39%) |  |
| **Hispanic – n (%)** | 10 (13%) | 19 (17%) | - | 14 (15%) | 44 (16%) | 0.7 |
| **TLC Global – mean (SD)** | 0.1 (0.3) | 0.4 (0.6) | 1.7 (1.1) | 1.3 (1.0) | 0.7 (1.0) | <0.001 |

Note: SD – standard deviation, TLC Global – Global score from the Scale for the Assessment of Thought Language and Communication.

### Clinical Ratings of Speech Disturbance

To clinically characterize language disturbances, all samples were given ratings using the 18 items from the Scale for the Assessment of Thought Language and Communication (TLC)^28^ and two additional speech-related items from the Scale for the Assessment of Negative Symptoms (SANS)^29^ which were not included in the TLC (SANS-06: Decreased Vocal Inflection; SANS-11: Increased Latency of Response). Consistent with previous reports,^21,28,30^ we found low prevalence (absent in >90% of the sample), low interrater reliability, or low sampling adequacy for six TLC items: echolalia, blocking, clanging, word approximation, self-reference, and stilted speech. These were not included in the analyses. Each of the remaining 14 items exhibited excellent interrater reliability (ICC³0.9).

### Speech Features

All samples were transcribed verbatim and processed through an automated pipeline to extract acoustic (prosody and voice quality, speaking tempo and pauses) and textual features (semantic distances, dysfluencies and speech errors, speech graph measures, lexical characteristics, sentiment, parts-of-speech, speech quantity). We initially selected 79 features for analysis. To improve interpretability of the graphs, we calculated variance inflation factors for each category of features and omitted redundant features. Twenty-seven features were included in the final analysis. Additional details are provided in the supplement.

### Clinical Assessments

A subset of participants was further characterized on psychosis symptoms, functioning, and select cognitive measures (n=125), which were included in the network analysis to provide context for the clinical speech factors and computational speech features. Clinical characteristics included measures of general psychosis symptoms, anxiety/depression, negative symptoms, verbal ability, social cognition, and functional impairment. Additional details are provided in the supplement.

### Factor Analyses

Exploratory factor analysis (eFA) was used to generate potential models for latent factors for clinical speech disturbance ratings in the cross-diagnostic sample. Usual assumptions were met: Bartlett Test of Sphericity: p<<0.001; Kaiser-Meyer-Olkin measure of sampling adequacy = 0.9; determinant = 0.0004. Visual inspection of the scree plot (Supplemental Figure 1) suggested 2-3 latent factors. We used the *psych* package^31^ in R^32^ to generate 2- and 3-factor principal axis solutions with promax rotations. We chose an oblique rotation because we hypothesized that latent factors may be correlated with one another. Confirmatory factor analysis (cFA) was used to examine fit statistics for the full cross-diagnostic sample and the SSD subgroup. Maximum likelihood estimation from *lavaan* package^33^ was used. The main group effect on factor scores was evaluated using ANOVA and pair-wise comparisons were made using t-tests.

### Network Generation

Speech factor scores were computed based on the final 3-factor model and a network was constructed to illustrate correlational relationships. Nodes represent factor scores, speech features, and clinical characteristics. Edges represent Spearman correlation coefficients with **ρ**>0.3 and p<0.01. The graph was plotted in R using the *igraph* package^34^ and network descriptors were calculated. All metadata, code for analysis, and resources for duplicating our factor score calculations are available at: https://github.com/STANG-lab/Analysis/tree/main/Factor-network.

## Results

### Latent Factors of Language Disturbance

Results of the eFA (Table 2, Supplemental Figures 2 & 3) suggested 2- and 3-factor models based on 14 clinical ratings for speech and language disturbance symptoms in the cross-diagnostic sample. The 2-factor model identified factors related to *disorganized speech* and *impaired expressivity* (decreased speech content and expressiveness), explaining 40% and 12% of the variance, respectively. The 3-factor model also produced the *impaired expressivity* factor (5% variance) and further divided disorganized speech into items consistent with *inefficient speech* (poor organization across ideas; 41% of variance) and *incoherent speech* (nonsensical or unintelligible utterances; 12% of variance). Each model was tested using cFA in both the overall sample and the SSD subgroup. The 2-factor model was a poor fit for both groups (Cross-diagnostic: Comparative Fit Index (CFI)=0.850, Tucker-Lewis Index (TLI)=0.821, Root Mean Square Error of Approximation (RMSEA)=0.095; SSD: CFI=0.827, TLI=0.793, RMSEA=0.118). Because the 3-factor model had multiple cross-loadings which are not suitable for cFA, we tested this model in two ways: first, by omitting the cross-loaded items (Poverty of Content of Speech, Derailment, and Loss of Goal) and then by including a separate fourth factor with the cross-loaded items. Both approaches demonstrated good fit for the final model in the cross-diagnostic sample: Without cross-loaded items (3-factor): CFI=0.965, TLI=0.953, RMSEA=0.047; With separate factor (4-factor): CFI=0.954, TLI=0.941, RMSEA=0.054. The fits were acceptable in the SSD subsample: Without cross-loaded items (3-factor): CFI=0.908, TLI=0.877, RMSEA=0.086; With separate factor (4-factor): CFI=0.917, TLI=0.893, RMSEA=0.085).

**Table 2.** Factor Loadings.

| 2-Factor Model |  | Speech and Language Items | 3-Factor Model |  |  |
| --- | --- | --- | --- | --- | --- |
| Disorganized Speech | Impaired Expressivity |  | Inefficient Speech | Incoherent Speech | Impaired Expressivity |
|  | 0.77 | Poverty of Speech (TLC-01) |  |  | 0.81 |
| 0.87 |  | Poverty of Content of Speech (TLC-02) | 0.46 | 0.47 |  |
| 0.65 | -0.34 | Pressured Speech (TLC-03) | 0.79 |  |  |
| 0.49 |  | Distractible Speech (TLC-04) | 0.58 |  |  |
| 0.85 |  | Tangentiality (TLC-05) | 0.92 |  |  |
| 0.89 |  | Derailment (TLC-06) | 0.58 | 0.39 |  |
| 0.65 |  | Incoherence (TLC-07) |  | 0.89 |  |
| 0.73 |  | Illogicality (TLC-08) |  | 0.67 |  |
| 0.38 |  | Neologism (TLC-10) |  | 0.58 |  |
| 0.72 |  | Circumstantiality (TLC-12) | 0.84 |  |  |
| 0.82 |  | Loss of Goal (TLC-13) | 0.45 | 0.43 |  |
| 0.66 |  | Perseverations (TLC-14) | 0.58 |  |  |
|  | 0.62 | Decreased Vocal Inflections (SANS-06) |  |  | 0.69 |
|  | 0.57 | Increased Latency of Response (SANS-11) |  |  | 0.56 |

Note: Loadings <0.3 are masked. TLC – Scale for the Assessment of Thought Language and Communication; SANS – Scale for the Assessment of Negative Symptoms.

### Group Differences in Factor Scores

Each of the three factors exhibited significant group effects (p<0.001) but with different patterns (Figure 1). Inefficient speech showed a graded effect with significant differences between each pair of groups; inefficient speech was highest in the SSD group, followed by PSY, then OD and HV. Incoherent speech was specific to psychosis and was elevated in both psychotic groups (SSD and PSY) compared to both nonpsychotic groups (OD and HV); with no significant difference between SSD and PSY or between OD and HV. Clinically significant impaired expressivity symptoms were present across all groups, but highest in the psychosis groups (SSD and PSY), intermediate for participants with other psychiatric disorders (OD), and lowest for healthy volunteers (HV).

**Figure 1.**
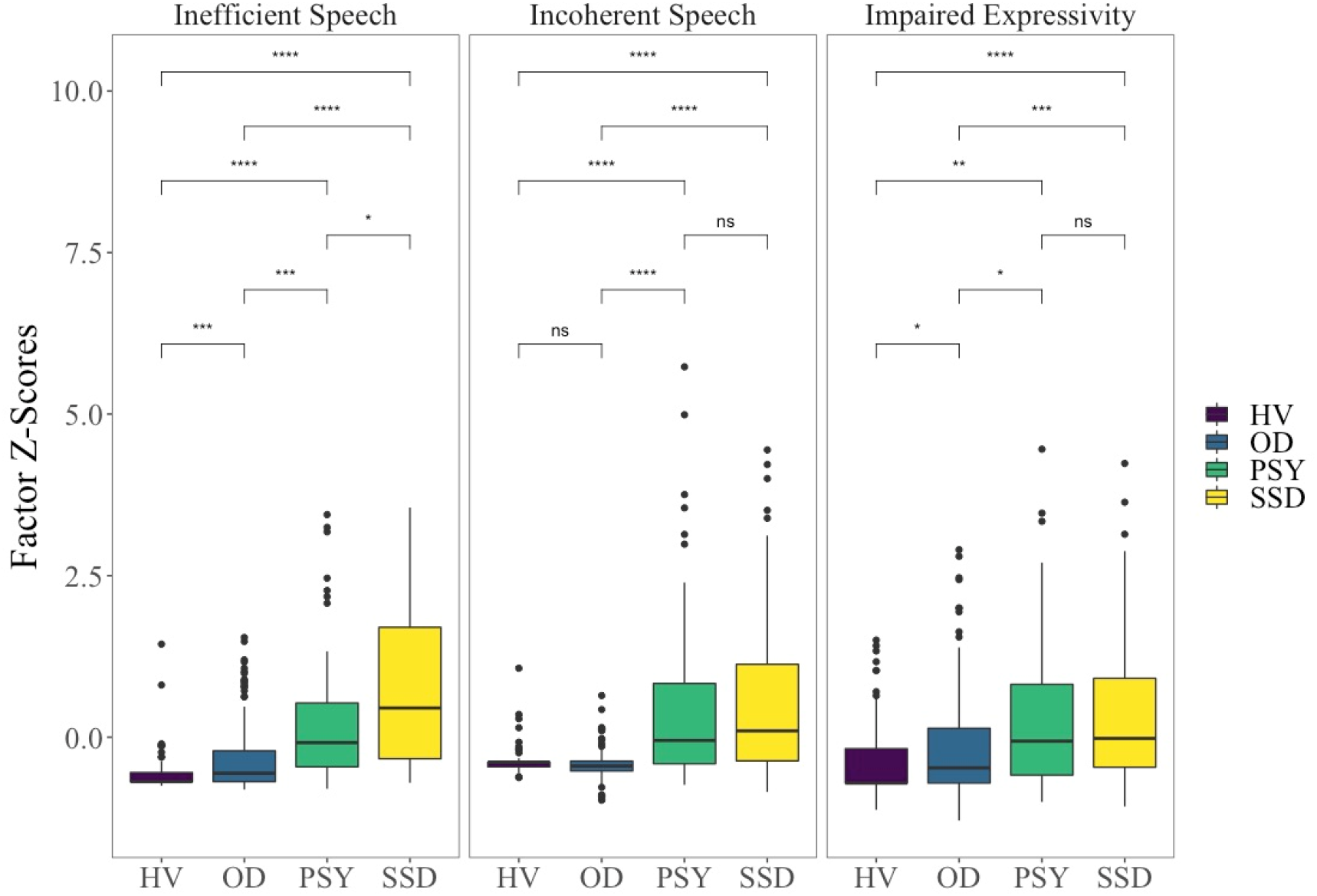
Group Differences In Factor Scores. Pairwise comparisons made using t-tests. *NS – Not significant; * p<0*.*05; ** p<0*.*01; ***p<0*.*001. HV – healthy volunteers; OD – other psychiatric disorders; PSY – other or undetermined psychotic disorders; SSD – schizophrenia spectrum disorders*.

### Relationships to Computational Features

Networks generated for the factor scores, computational language features, and clinical symptoms also showed distinct relationships with each of the three factors, as well as different patterns for the cross-diagnostic (Figure 2) versus SSD samples (Figure 3). For the cross-diagnostic group, all clinical characteristics were tightly interrelated. The overall density was 0.19. Impaired expressivity was the most highly connected speech factor (Degree (D)= 14; Betweenness Centrality (BC)=181.4), followed by inefficient speech (D=13, BC=8.5) and then incoherent speech (D=10, BC=3.7). For the SSD subgroup, overall density was 0.09, reflecting greater separation among clinical characteristics and speech features. Inefficient speech was the most highly connected (D=9, BC=122.3), followed by impaired expressivity (D=7, BC=64.8), and then incoherent speech (D=6, BC=13.9).

**Figure 2.**
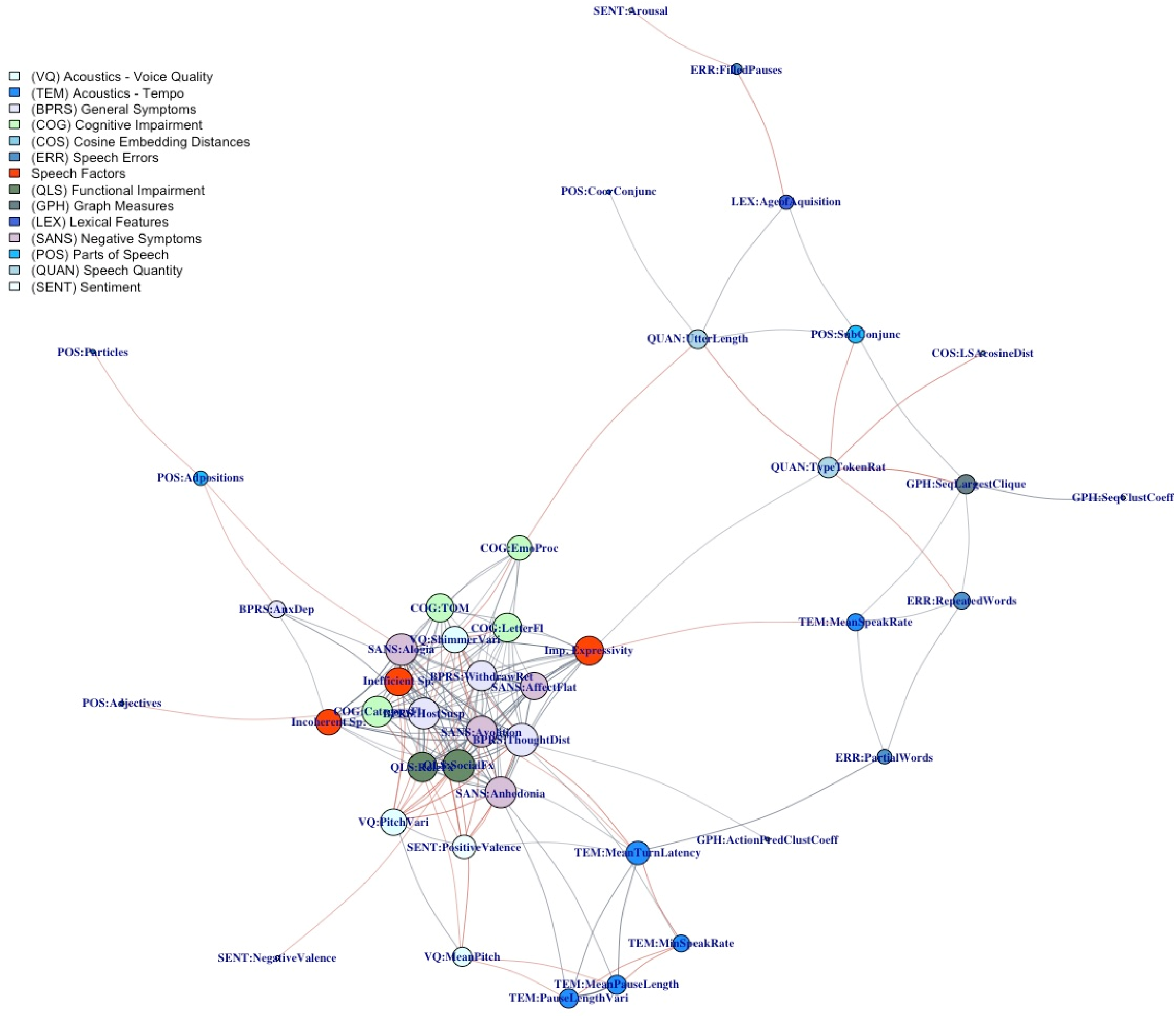
Cross-Diagnostic Factors, Features, and Symptoms Network: Nodes represent factor scores, computational acoustics and lexical features, and clinical symptoms ratings and scores; size is proportional to the degree of the node. Edges represent Spearman correlation coefficients with cutoff of **ρ**=0.3 and p=0.01; weight is proportional to absolute value; gray edges are positive correlations and red edges are negative correlations.

**Figure 3.**
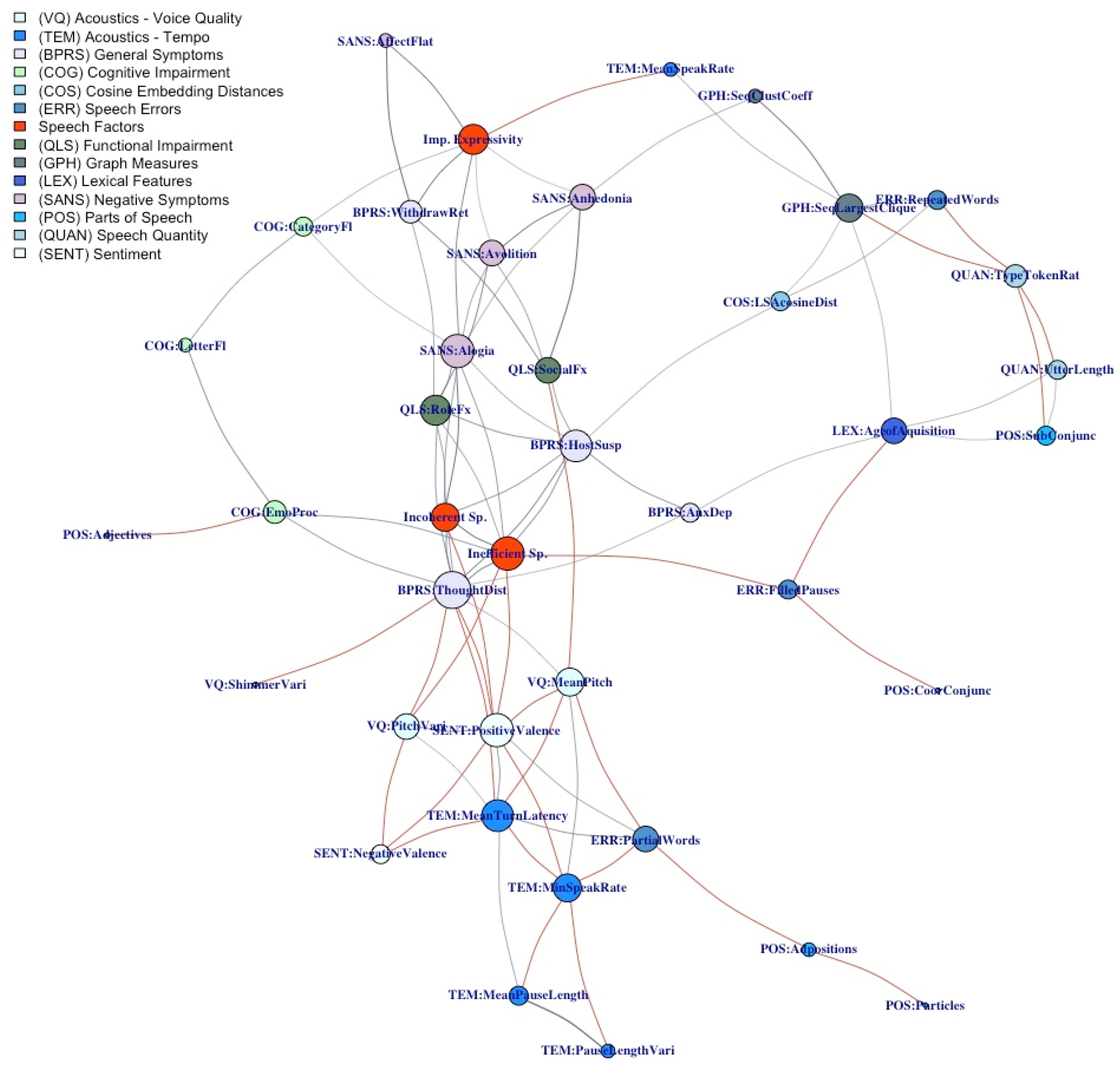
Schizophrenia Spectrum Disorders Subgroup – Factors, Features, and Symptoms Network: Nodes represent factor scores, computational acoustics and lexical features, and clinical symptoms ratings and scores; size is proportional to the degree of the node. Edges represent Spearman correlation coefficients with cutoff of **ρ**=0.3 and p=0.01; weight is proportional to absolute value; gray edges are positive correlations and red edges are negative correlations.

For both groups, the three speech factors had distinct patterns of correlations with other features. Among SSD, inefficient speech was most closely correlated with incoherent speech, repeated words, higher speaking rate, and impairment in emotion processing; other significant relationships to computational features included largest clique size in sequential graphs, cosine embedding distances using latent semantic analysis, and higher age of acquisition of words. Incoherent speech was most closely correlated with impaired theory of mind, inefficient speech, alogia, and impaired emption processing; other significant relationships included higher speaking rate, higher arousal sentiment, greater use of subordinate conjunctions, and decreased variance in vocal shimmer. Impaired expressivity was most tightly connected to withdrawal/retardation symptoms, affective flattening, alogia, and impairment in interpersonal functioning; other significant relationships included decreased mean pitch, decreased clustering coefficient in action-predication graphs, and decreased variance in shimmer.

## Discussion

In this study, we identified a 3-factor model that describes speech and language disturbance with good fit in cross-diagnostic and SSD samples. An *impaired expressivity* factor included items related to decreased quantity and expressiveness. Two interrelated disorganized factors emerged: an *inefficiency factor* included items describing poorly-related, redundant, or excessive speech, and an *incoherent factor* included items relating to nonsensical or unintelligible speech. Fittingly, derailment, loss of goal, and poverty of content of speech were cross-loaded on both the inefficient and the incoherent factors. Our confirmatory models suggest a good fit for the 3-factor model in the cross-diagnostic sample, and adequate fit for the SSD subgroup which is comparable to previously reported models.^21^ The distinction between impaired expressivity and disorganization-type symptoms is well-replicated in SSD^21^ and cross-diagnostically.^2^ Specifically, a latent factor for poverty of thought and decreased expressiveness has been found by other groups and using other rating scales.^35,36^ Factor analysis of the Thought and Language Index also suggests that unusual word usage, sentence structure, and logic (consistent with our *incoherence* factor) may be differentiated from distractibility and perseveration (included in our *inefficiency* factor).^37^ Overall, our model is very similar to the 3-factor model for formal thought disorder reported by Stein et al. in their cross-diagnostic sample of N=1,071.^25^ They describe an emptiness factor (poverty of speech and content, increased latency, and blocking), a disorganization factor (tangentiality, circumstantiality, derailment, pressure of speech), and an incoherence factor (incoherence, illogicality, distractibility). The principal differences are that we include decreased vocal inflection in our impaired expressivity factor (since we are looking at speech as a whole, and not specifically thought disorder), and that we prefer the term “inefficiency” over “disorganization” because “disorganization” is an overly broad term than can include incoherence and other items. Notably, Stein et al. report distinct correlates to brain structure for their three factors. The similarity between our findings, and the fact that the samples were collected in different languages and rated by different teams, provides added confidence for the 3-factor model we propose.

The 3-factor model proposed here can be understood in the context of linguistic theories on pragmatics. In classical Gricean pragmatics, a speech act is carried out successfully when the addressee is made to recognize the speaker’s communicative intent (“Meaning-intention” or “M-intention”).^38,39^ Understanding the M-intention necessitates not only deciphering the semantic content (literal meaning) of what is spoken, but also the conversational implicatures which are grounded in the context of the discourse and in the cooperative principle. Per Grice, the cooperative principle assumes that the speaker is always following four maxims: (1) Quantity: be informative, but not overly informative; (2) Quality: be truthful, to the best of one’s knowledge; (3) Relation: be relevant; and (4) Manner: be perspicuous (clear). Along these lines, we can characterize the impaired expressivity factor as violations of the maxim on quantity—specifically, delivery of too little information. Inefficiency and incoherence both involve violations of relation, manner, and (excessive) quantity, but differ in degree. With incoherent speech, the violations are severe to the point of preventing the M-intention from being inferred. With inefficient speech, the violations of the cooperative principle are perceived, but the M-intention can still be ascertained. We can also differentiate between incoherent and inefficient disorganized speech by referring to Grosz’s theories on centering and discourse structure.^40,41^ She suggests that, just as individual sentences can be broken down into a hierarchical structure of phrases which give meaning to one another, so can discourse be broken into a hierarchy of discourse segments (each containing one or more utterances). In coherent discourse, individual utterances within a discourse segment collaborate to convey the discourse segment purpose (local coherence), and multiple discourse segments relate to one another to satisfy the overall discourse purpose (global coherence). With respect to our work, we would characterize inefficiency as disruptions in global coherence, where there are disruptions or inefficiencies in satisfying the overall discourse purpose, while incoherence arises from disruptions to local coherence that cause the discourse segment purpose to be obscured.

We also found distinct and significant diagnosis group effects for each of the three factors, suggesting that the model is not just statistically and theoretically sound, but also clinically meaningful. There were intermediate effects for nonpsychotic disorders in inefficiency and impaired expressivity, but incoherence was specific to people with psychosis and rarely elevated in either healthy volunteers or people with nonpsychotic disorders. Intuitively, this pattern may be explained by the sensitivity of impaired expressivity and inefficiency to non-psychiatric (e.g., personality, culture, social context) and non-specific factors (e.g., impaired attention, psychomotor retardation, ruminations and repetitive thinking). In contrast, incoherence may be more specific to psychosis-related brain-changes. This hypothesis should be further evaluated.

There were notable differences between the cross-diagnostic and SSD groups in how factor scores related to other clinical characteristics and quantitative speech features. In the cross-diagnostic sample, all clinical characteristics were tightly interrelated, likely representing a common dimension of psychopathology. Impaired expressivity showed the highest betweenness centrality and served as a bridge between clinical characteristics and quantitative speech features. In the SSD subsample, as expected, impaired expressivity was more closely related to negative symptoms like affective flattening, withdrawal, anhedonia, and avolition, while incoherent and inefficient speech were more closely related to positive symptoms like thought disturbance, hostility and suspiciousness. Among SSD, inefficient speech had the highest betweenness centrality, and connected multiple symptom dimensions, functioning, and quantitative speech features. For both groups, each speech factor was closely related to other clinical characteristics but demonstrated distinct relationships with quantitative speech features. Some of the relationships illustrated in our network diagrams reflect those reported in previous works. For example, disorganized speech has been connected with poor functioning and greater cognitive impairment.^42^ Affective flattening and anhedonia have been related to decreased speaking rate.^43,44^ In general, we interpret the results from our network analyses to suggest that each speech factor taps consistently into critical clinical phenomena, but that computational modeling should be pursued separately for the individual factors, and possibly separately for different clinical populations.

There were several limitations in our work which should be addressed in future studies. There were heterogeneities in the data collection methods. The network analysis does not account for moderating or confounding variables, such as gender, age, and socioeconomic status. All samples were collected in English and rated with the TLC scale (with 2 SANS items) at a single site. A larger multi-lingual study using multiple rating scales would further support the generalizability of our findings. The fit of the 3-factor model for other (nonpsychotic) diagnostic groups or within individual SSD diagnoses was not examined because it was outside the scope of this paper. Furthermore, both clinical ratings and quantitative speech features may be affected by the context of the interview and the nature of the task. There may also be group by context effects on speech. The performance of predictive models may be further improved by standardizing and accounting for these variations.

In this study, we report a cross-diagnostic 3-factor model for speech and language disturbance which is supported by good statistical measures, intuitive, applicable to SSD, and relatable to linguistic theories. *Impaired expressivity, inefficiency*, and *incoherence* show meaningfully distinct patterns in different diagnostic groups, with incoherent speech being most specific to psychosis. Each factor was closely related to a network of other clinical characteristics but demonstrated distinct relationships to quantitative speech features. The factors also intuitively inspire different computational strategies. In conclusion, the 3-factor model reported here is a valuable framework for understanding speech and language disturbance cross-diagnostically and in SSD particularly, and the factor scores are appropriate targets for modeling with quantitative speech features.

## Data Availability

All metadata, code for analysis, and resources for duplicating our factor score calculations are available at: https://github.com/STANG-lab/Analysis/tree/main/Factor-network

## Acknowledgements

This project was supported by the Brain and Behavior Research Foundation Young Investigator Award (SXT) and the American Society of Clinical Psychopharmacology Early Career Research Award (SXT). Data for a portion of the participants (n=210) was collected in partnership with, and with financial support from, Winterlight Labs, Inc, but the conceptualization for this project, computation of speech features, and analyses were completed independently. We thank the participants for their contributions. We also thank Danielle DeSouza, Bill Simpson, Jessica Robin, and Liam Kaufman of Winterlight Labs for their ongoing collaboration. We are grateful to the clinicians and leaders at Zucker Hillside Hospital for their support of our work, including Drs. Michael Birnbaum, Anna Costakis, Ema Saito, Anil Malhotra, and John Kane. We thank James Fiumara and Jonathan Wright from the Linguistic Data Consortium, and Aamina Dhar, Grace Serpe, Jessica Guo and Styliana Maimos for their help in the transcription process.

