## Supplemental Materials for "Latent Factors of Language Disturbance and Relationships to Quantitative Speech Features"

### Methods:

*Participants*

Of the 343 speech samples analyzed in this study:

- 47 samples of free speech were collected from publicly-available educational videos of psychiatric interviews on YouTube, including 28 samples from people designated as having a psychotic disorder, and 19 samples from people reported to have a non-psychotic psychiatric condition (further detailed by Krell et al.[1]). Audio samples were cut from posted interviews to standardize length (1.5-2min) and isolate open-ended narratives, consistent with the other speech samples. Because psychotic disorder diagnosis could not be confirmed, samples from individuals identified as having psychotic disorders were designated as unconfirmed but probably psychosis (PSY). Analysis of these samples were done on publicly available data, so this portion of the study was not considered human subjects research and did not undergo IRB review.
- The remaining 296 speech samples were collected from participants enrolled in ongoing studies of speech biomarkers in psychiatric disorders. Participants were recruited from inpatient and outpatient programs at the Zucker Hillside Hospital, rosters of past research participants, and posts on the internet. Free speech was recorded in response to two open-ended questions asking for a self-description and report on recent experiences. Of the 296 speech samples, 153 were collected in-person via a proprietary iOS app developed by Winterlight Labs, 17 were collected in-person via a standard digital voice recorder, and 126 were collected virtually over Microsoft Teams. These samples were collected as a part of larger studies which included additional language tasks (picture descriptions, fluency tasks) and clinical assessments. TLC and SANS ratings were given holistically for the overall assessment.

*Speech Features*

Processing of all 343 samples occurred through an automated pipeline. We initially selected 79 features for evaluation. 52 features were omitted after VIF analysis due to redundancy with other features. A cutoff of VIF<5 was used for all categories except for cosine embedding distances, where we used VIF<10 because no features had a VIF<5. See Supplemental Table 1 for final feature table (n=27).

- Transcription: Human annotators produced verbatim transcriptions, including speaker labels and timestamps. Utterance boundaries were defined intuitively based on pauses and syntax. Dysfluencies and speech errors such as filled pauses, partial words and repeated words were tagged.
- Prosody and voice quality features were extracted from OpenSMILE[2] using the interspeech13 configuration.[3] Recordings were diarized for participant vs. interviewer speech and only participant segments were used to calculate prosodic and voice quality features. Starting features = 6.
- Speaking tempo and pauses were calculated by first aligning transcripts with audio recordings using the Montreal forced aligner.[4] Segments were labeled as voiced vs. unvoiced, participant vs. interviewer. Pause lengths and rate of speech were quantified for the participant. Starting features = 7.
- Semantic cosine embedding distances were calculated by first removing filled pauses, partial words, repetitions, and NLTK stop words.[5] Then, word embeddings were calculated using Latent Semantic Analysis (LSA),[6] Word2Vec,[7] and Glove.[8] Utterance embeddings were calculated by taking the simple mean of the word embeddings, or using TF-IDF weighting.[9] Starting features = 6.
- Speech errors and dysfluencies were tagged during transcription, counted, and standardized by the total word count. Starting features = 3.
- Speech graph features were generated using sequential, cooccurrence and action-predication methods further detailed by Nikzad et al.[10] Briefly, sequential graphs were created by connecting each word to the next, cooccurrence graphs were made by connecting all words that appear in each sentence together, and action-predication graphs were produced by connecting entities that act upon each other and the content of an activity to its participants. The graph features describing network size (number of nodes, number of edges, average shortest path length, and network diameter) and network connectedness (average degree, average weighted degree, graph density, average clustering coefficient, size of the largest clique, size of the largest strongly connected component and number of triangles) were calculated for each graph type Starting features = 31.
- Lexical characteristics for age of acquisition,[11] prevalence,[11] and semantic diversity[12] were calculated based on published norms. Starting features = 3.

- Sentiment features examined valence, arousal, and dominance characteristics of words based on published norms.[13] Starting features = 5.
- Parts-of-speech were counts of POS tags generated using spAcy,[14] standardized by the total word count in the sample. Starting features = 14.
- Speech quantity measures such as word count, utterance count, average utterance length, were directly calculated from the transcripts. Starting features = 4.

#### Supplemental Table 1: Speech Features

| Category | Feature | Description |
|---|---|---|
| COS: Semantic Cosine Embedding Distances | LSAcosineDist | Utterance embeddings calculated from mean embeddings of individual words. Cosine distance calculated between adjacent sentences. |
| ERR: Speech Errors and Dysfluencies | FilledPauses | E.g., um, uh, er; count per 100 words. |
| ERR: Speech Errors and Dysfluencies | PartialWords | Dysfluent words which were interrupted by the speaker and not completely uttered. E.g., "I went to the **st-** store." |
| ERR: Speech Errors and Dysfluencies | RepeatedWords | Dysfluent repeated words or segments. E.g., "I **I** went to the store." "I went **I went** to the store." |
| GPH: Graph Features | ActionPredClustCoeff | Clustering coefficient calculated from action-predication graph with dysfluencies, repetitions removed |
| GPH: Graph Features | SeqClustCoeff | Clustering coefficient calculated from sequential graph with dysfluencies, repetitions removed |
| GPH: Graph Features | SeqLargestClique | Largest clique (interconnected component) calculated from sequential graph with dysfluencies, repetitions removed |
| LEX: Lexical Characteristics | AgeofAquisition | Average age at which word is acquired |
| POS: Parts-of-speech | Adjectives | After removing repeated segments, count of adjectives per 100 words |
| POS: Parts-of-speech | Adpositions | After removing repeated segments, count of prepositions per 100 words |
| POS: Parts-of-speech | CoorConjunc | After removing repeated segments, count of coordinating conjunctions per 100 words |
| POS: Parts-of-speech | Determiners | After removing repeated segments, count of determiners per 100 words |
| POS: Parts-of-speech | Particles | After removing repeated segments, count of particles per 100 words |
| POS: Parts-of-speech | SubConjunc | After removing repeated segments, count of subordinating conjunctions per 100 words |
| QUAN: Speech Quantity | TypeTokenRat | Ratio of unique tokens to total number of tokens |
| QUAN: Speech Quantity | UtterLength | Mean length (in words) of each utterance in the verbatim transcription |
| SENT: Sentiment | Arousal | Degree to which words reflected arousal state (+) vs. calm (-) |
| SENT: Sentiment | NegativeValence | Degree of negative valence expressed; mean valence score of all negative-valence-coded content words |
| SENT: Sentiment | PositiveValence | Degree of positive valence expressed; mean valence score of all positive-valence-coded content words |
| TEM: Tempo and Pauses | MeanPauseLength | Mean duration of pauses in participant speech |
| TEM: Tempo and Pauses | MeanSpeakRate | Words per utterance divided by utterance duration; mean across all participant speech. |
| TEM: Tempo and Pauses | MeanTurnLatency | Mean initial pause duration for participant turns (after interviewer speech) |
| TEM: Tempo and Pauses | MinSpeakRate | Words per utterance divided by utterance duration; minimum value across all participant speech. |
| TEM: Tempo and Pauses | PauseLengthVari | Standard deviation of pause lengths |
| VQ: Voice Quality and Prosody | MeanPitch | Mean of F0 |
| VQ: Voice Quality and Prosody | PitchVari | Standard deviation of F0 |
| VQ: Voice Quality and Prosody | ShimmerVari | Standard deviation of the local (frame-to-frame) shimmer (amplitude deviations between pitch periods) |

*Clinical Assessments*
A cross-diagnostic subset of participants (n=125) were assessed with the following clinical rating scales. To generate consistent directionality, the functioning and cognitive scores were inverted so that higher scores reflected greater impairment, as for the symptom ratings.

- BPRS: Brief Psychiatric Rating Scale,[15] with 5 factors calculated as per Overall et al.
- SANS: Scale for the Assessment of Negative Symptoms,[16] with global scores for 4 domains as per Andreasen.
- QLS: Heinrich's Quality of Life Scale.[17]
- Cognitive Impairment (COG): Social cognition was assessed with the Hinting Task,[18] Penn emotion recognition task,[19] and verbal fluency tasks (F-letter, and category).

**Supplemental Table 2: Clinical Characteristics**

| Category | Item | Description |
|---|---|---|
| BPRS | AnxDep | Factor score for anxious depression: Anxiety, Guilty Feelings, Depressive Mood |
| BPRS | HostSusp | Factor score for hostile suspiciousness: Hostility, Suspiciousness, Uncooperativeness |
| BPRS | ThoughtDist | Factor score for thought disturbance: Conceptual Disorganization, Hallucinatory Behavior, Unusual Thought Content |
| BPRS | WithdrawRet | Factor score for withdrawal retardation: Emotional withdrawal, Motor retardation, Blunted affect |
| SANS | AffectFlat | Global score for affective flattening |
| SANS | Alogia | Global score for alogia |
| SANS | Avolition | Global score for avolition |
| SANS | Anhedonia | Global score for anhedonia and asociality |
| QLS | RoleFx | (Inverted) Subscore for role functioning |
| QLS | SocialFx | (Inverted) Subscore for social functioning |
| COG | TOM | (Inverted) Hinting score |
| COG | EmoProc | (Inverted) Penn emotion recognition score |
| COG | LetterFl | (Inverted) Performance on F-letter fluency task |
| COG | CategoryFl | (Inverted) Performance on category (animals) fluency task |

*Factor Analyses*

**Supplemental Figure 1: Scree Plot for Exploratory Factor Analysis**

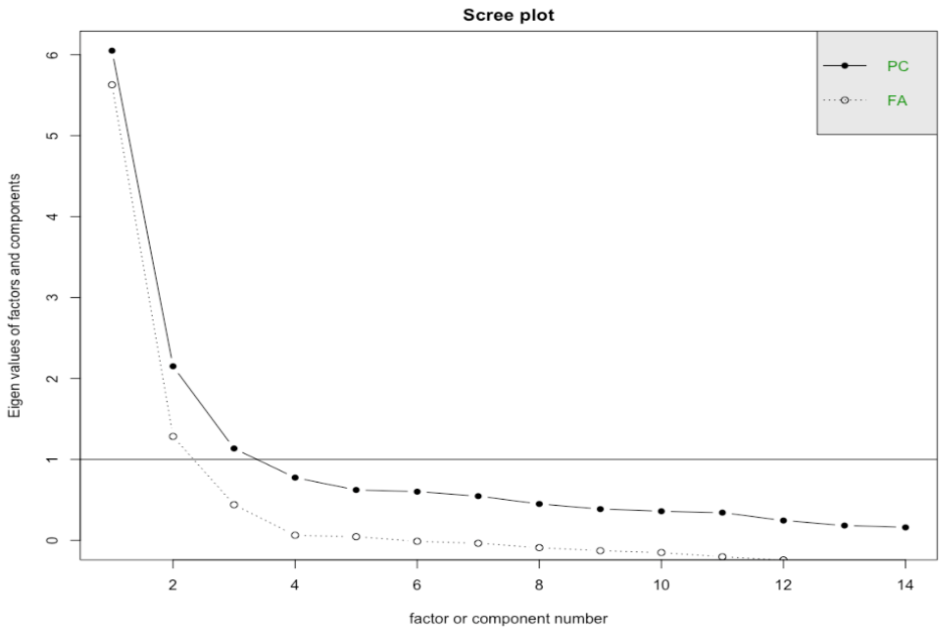

**Supplemental Figure 2: 2-Factor Model**

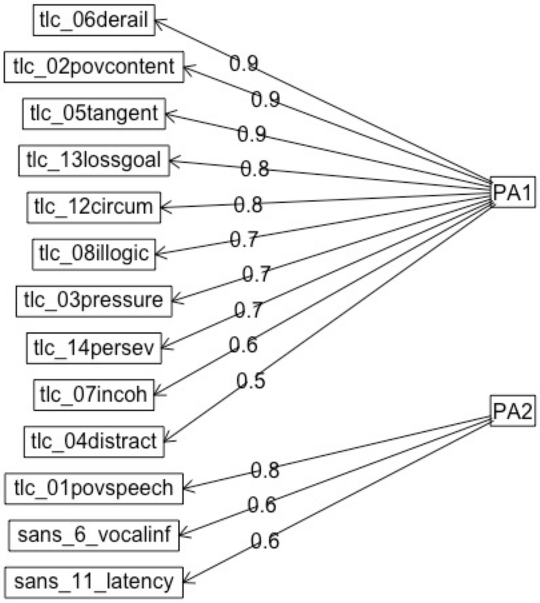


**Supplemental Figure 3: 3-Factor Model**

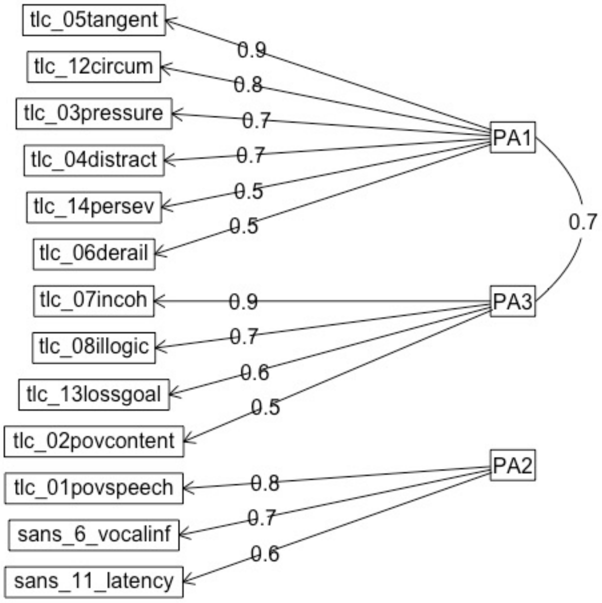
